# Patterns of convergence and divergence between bipolar disorder type I and type II: evidence from integrative genomic analyses

**DOI:** 10.1101/2021.12.15.21267813

**Authors:** Yunqi Huang, Yunjia Liu, Yulu Wu, Yiguo Tang, Siyi Liu, Liling Xiao, Mengting Zhang, Shiwan Tao, Min Xie, Minhan Dai, Mingli Li, Hongsheng Gui, Qiang Wang

## Abstract

Genome-wide association studies (GWAS) analyses have revealed genetic evidence of bipolar disorder (BD), but little is known about genetic structure of BD subtypes. We aimed to investigate genetic overlap and distinction of bipolar type I (BDI) & type II (BDII) by conducting integrative post-GWAS analyses. This study utilized single nucleotide polymorphism (SNP)-level approaches to uncover correlated and distinct genetic loci. Transcriptome-wide association analyses (TWAS) were then approached to pinpoint functional genes expressed in specific brain tissues and blood. Next, we performed cross-phenotype analysis including exploring the potential causal associations between BDI & II and drug responses and comparing the difference of genetic structures among four different psychiatric traits. Our results find SNP-level evidence revealed three genomic loci, *SLC25A17, ZNF184* and *RPL10AP3* shared by BDI & II, while one locus (i.e., *MAD1L1)* and significant gene sets involved in calcium channel activity, neural and synapsed signals that distinguished two subtypes. TWAS data implicated different genes effecting BDI & II through expression in specific brain regions (e.g., nucleus accumbens for BDI). Cross-phenotype analyses indicated that BDI & II share continuous genetic structures with schizophrenia (SCZ) and major depression disorder (MDD), which help fill the gaps left by the dichotomy of mental disorder. These combined evidences illustrate genetic convergence and divergence between BDI & II and provide an underlying biological and trans-diagnostic insight into major psychiatric disorders.

## Introduction

Bipolar disorder (BD) is one of the most severe psychiatric disorders, characterized by mood states fluctuation. As one of the top causes of disability worldwide, BD affects more than 40 million people worldwide with the lifespan prevalence among 1%∼4%^1, 2^, early onset in adolescents, and elevated risk of suicide^3^.

Population and molecular studies have proved evidence into the complex etiology of BD. Twin and family studies have estimated that the heritability of BD is over 70%^4, 5^ GWAS have brought deeper insights to BD^6, 7^ compared with previous population genetics studies^8-10^. The largest-scale GWAS of BD was recently processed by Psychiatric Genomic Consortium Bipolar Disorder Working Group (PGC3 BD)^11^ and 64 genome-wide significant loci were identified. However, it failed to display increasing of single nucleotide polymorphism (SNP)-level heritability (h^2^_SNP_) of BD11, 12.

BD can be categorized into several major subtypes: BD type I (BDI) and type II (BDII), cyclothymia, and other specified bipolar and related disorder, according to the Diagnostic and Statistical Manual Disorders, Fifth Edition (DSM-5)^8^. BDI requires manic episodes at least once despite depression states, and BDII, is defined as more than one depressive and hypomanic state. The lifetime prevalence of BDI (0.4%-1.2%)^1, 2, 13^ differs from BDII (0.1%-2.5%)^1, 2, 13^ in worldwide. In addition, clinical presentations and severity vary in two subtypes^1, 14^, however, their biological differences remained unclear due to insufficient sample size or substandard clinical classification^15, 16^.

Under the dichotomic diagnostic system, BD is hard to be distinguished from schizophrenia (SCZ), and major depression disorder (MDD). BD with psychotic symptoms or manic BD performs and behaves similarly as SCZ; and oppositely, depressive BD is always misdiagnosed as MDD, resulting in limited therapeutic effects. Additionally, the second-generation antipsychotics play an important role in treatment of BD, SCZ and MDD, indicating potential shared biological mechanism among these phenotypes. Linkage disequilibrium (LD) score analysis indicated that BDI is much more genetically correlated with SCZ, whereas the genetic correlation of BDII with MDD is higher^11^. These provide a new perspective into genetic correlation between BDI, BDII, SCZ and MDD. Moreover, molecular genetic studies have uncovered overlapped risk factors between genomic architecture of psychiatric disorders^6, 11, 12, 15^; however, current diagnostic systems failed to elucidate it clearly.

To better understand BD etiology and taxonomy, here our study aimed to provide more evidence through post-GWAS analyses. Integrative Omics approaches were adopted to navigate functional genes expressed in the influenced brain regions. Additionally, we explored cross-phenotype genetic structure in adult psychiatric disorders. We are challenging to enrich the research domain criteria (RDoC), re-evaluate and provide new evidence for the cross-disease diagnosis of mental disorders.

## Methods and Materials

### Study Design and GWAS data resources

The genome-wide association (GWA) meta-analysis summary data of BDI and II were from the Psychiatric Genomics Consortium Bipolar Disorder Working Group (PGC3 BD), containing BDI (25,060 cases and 449,978 controls) and BDII (6,781 cases, 364,075 controls), respectively. All participants are European descents, and diagnosis is explicitly based on international consensus criteria (DSM-IV, ICD-9 or ICD-10). Details on participant and cohort information, quality control can be accessed11. Nominal significant instrumental variants (IVs) of response to lithium salt in bipolar disorder^17-19^ were downloaded from National Human Genome Research Institute– European Bioinformatics Institute (NHGRI-EBI) GWAS Catalog^20^ (https://www.ebi.ac.uk/). The latest and biggest GWA meta-analysis summary statistics for SCZ (69,369 cases; 236,642controls)^21^, and MDD except samples in 23andMe dataset (59,851 cases and 113,154 controls)^22^ were also downloaded from PGC website. Overall post-GWAS analysis pipeline is shown in Figure 1.

**Figure 1.**
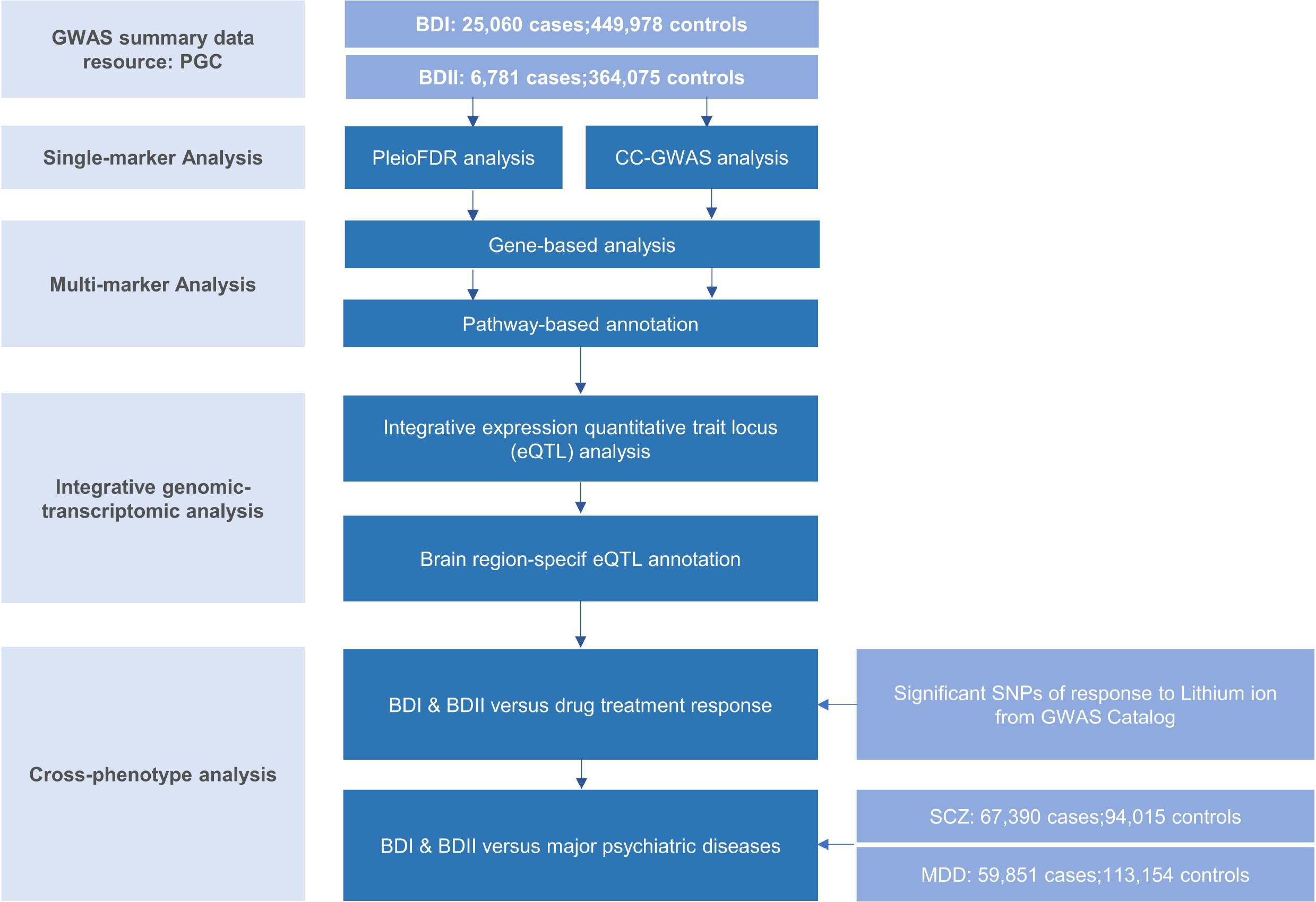
Workflow of key methodological steps in this study. PGC: psychiatric genomics consortium, BDI: bipolar disorder type I, BDII: bipolar disorder type II, SCZ: schizophrenia, MDD: major depression disorder.

### Single-marker Analysis

#### PleioFDR analysis

For genetic overlap, we used pleiotropy-informed conditional false discovery rate (pleioFDR) methods^23^, including conditional FDR (condFDR), an extension of the standard FDR, and conjunctional FDR (conjFDR) analysis, defined in turn as the maximum of the two condFDR values. The pleioFDR provided a conservative estimate of the FDR associated with both phenotypes, and was applied to identify specific shared loci. Based on an empirical Bayesian statistical framework, this statistical framework increased statistical power in detecting SNPs that did not reach genome-wide significance. Independent significant SNPs was defined with condFDR < 0.01.

#### CC-GWAS analysis

For genetic uniqueness, we applied CC-GWAS method^24^. CC-GWAS perceives differences in minor allele frequencies (MAF) across two traits by analyzing case-control GWAS summary statistics for each other. It weighted the effect size using two methods, ordinary least squares (OLS) weights and Exact weights. To effectively control an increased type I error rate caused by suspect null-null SNPs, SNPs with p_OLS_ <5×10^−8^ were remained. Then those failed to pass the required level of significance of CC-GWAS (p_EXACT_ >1×10^−4^) were excluded to avoid type I error rate caused by suspect stress test SNPs.

### Multi-marker Analysis

#### Gene-based analysis

Independent genomic loci were mapped by shared and trait-specific SNPs from GWAS summary data of BD I, BD II and CC-GWAS results using ANNOVAR employed in Functional Mapping and Annotation of GWAS (FUMA)^25^ online platform (https://fuma.ctglab.nl/). Significant SNPs were firstly selected by LD r^2^□>□0.6 within a 10kb windows. Second, we narrowed lead SNPs with LD r^2^□>□0.1 with the same window. Genomic risk loci were identified by merging lead SNPs if they were closer than 250 kb, thus containing multiple lead SNPs. The European samples retrieved from the phase 3 of the 1000 Genomes Project (1000G EUR) ^26^ was used to calculate LD.

To further define independent genomic loci diverged in BDI & II, we utilized MAGMA v1.6 implemented in FUMA^25^. Gene locations and boundaries were from the NCBI Build GRCh37 assembly. A locus that was not reported by original GWAS or GWAS Catalog was considered as novel.

#### Pathway-based annotation

Functional annotation was performed to uncover the likely biological mechanisms linking and distinguishing BDI & II. Enrichment for the genes mapped to all (candidate and lead) SNPs and genes nearest to lead SNPs in the identified shared loci was evaluated by the Molecular Signatures Database (MsigdB) via a hypergeometric test implemented in FUMA^25^. Genes without unique entrez ID were excluded. Pathway containing less than 2 genes was removed. The results were adjusted by Benjamini-Hochberg false discovery rate (BH FDR) of 0.05.

### Integrative genomic-transcriptomic analysis

#### Integrative expression quantitative trait locus (eQTL) analysis

To detect important but non-genome-wide significant sites, we firstly used summary-data-based Mendelian randomization (SMR)^27^ to estimate loci with strong evidence of causal effects of brain^28-31^ and blood^32^ via gene expression in BDI and BDII risk. SMR analysis was limited to significant cis eQTL (p_eQTL_ < 5×10^−8^), with minor allele frequency (MAF) > 0.20, and passing Heterogeneity In Dependent Instruments outlier (HEIDI-outlier) test (p≥0.01) due to its conservativeness^27, 33^. Significant loci were filtered after multiple testing and within 1MB distance from each probe.

#### Brain region-specific eQTL annotation

Likewise, we conducted brain-specific analyses using e-MAGMA^34^ and FUSION^35^, a transcriptome-wide association study (TWAS) to map genes based on precomputed tissue-specific eQTL statistics leveraging 13 brain tissues of GTEx v8^36^ and test whether SNPs influencing gene expression are also associated with BDI & II. The 1000G EUR were used as reference dataset to account for LD between SNPs. FDR correction were also applied to control the multiple tests performed on the numbers of genes in each process.

### Cross-phenotype analysis

#### BDI & II versus lithium treatment responses

We then investigate bi-directional causal relationships between BDI & II and lithium salt response using the “TwoSampleMR” (Version 0.5.3) and “MendelianRandomization” (Version 0.5.1) R packages. We selected independent SNPs with P-value < 5×10^−6^ and harmonized exposure and outcome data for both directions. For two sample MR analysis, we used inverse variance weighted (IVW), Weighted median based regression and MR-Egger as the primary method. MR-Egger intercept test and MR pleiotropy residual sum and outlier (MR-PRESSO) test ^37^ were used to evaluate potential IV horizontal pleiotropy.

#### BDI & II versus major psychiatric diseases

To explore genetic causal associations between BDI, BDII, SCZ, and MDD, we conducted bi-directional MR analysis for four traits to generate hypotheses concerning the direction of causation between each pair of the heritable variables using generalized Summary-data-based Mendelian Randomization (GSMR)^38^. We denoted x as an exposure trait significantly associated with z (Genetic variants), a putative causal trait for y (the outcome trait) for the same z. The effect of x on y was calculated by two-step least squares approach. The estimate 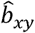 and its standard error from multiple Instrumental variants were associated with the exposure trait at a genome-wide significant level (p < 5×10^−8^). Attribute to insufficient instruments included in analyses, a p-value threshold of 5×10^−5^ was used. In GSMR, genetic instruments with pleiotropic effects are detected and eliminated by the HEIDI-outlier procedure, which was the same with SMR. We used default options in GSMR with HEIDI testing for the detection of instrumental outliers (LD r^2^ < 0.05, and at least 10 SNPs were required).

To estimate the genetic correlation between the two bipolar subtypes, SCZ and MDD, we applied MiXeR^39, 40^ as a polygenic overlap analysis. Univariate models estimated polygenicity (estimated number of variants) and discoverability (the average magnitude of additive genetic associations across variants) of each phenotype. Bivariate Gaussian mixture models were also applied to estimate the number of variants influencing each trait that explained 90% of h^2^snp and their overlap between each other. MiXeR calculated a Dice coefficient, a ratio of shared variants to the total number of variants, to evaluate the polygenic overlap. In line with Akaike information criterion (AIC), MiXeR evaluated model fitting based on current power of input summary statistics.

## Results

### Genetic overlaps between bipolar type I and II

For signals shared by BDI and BDII, the pleioFDR analysis identified 74 significant SNPs (p<0.01) that are mapped to 3 genomic loci (Table 1 and Figure 2A): zinc finger protein 184, *ZNF184*, mapped by *rs67240003* (p_FDR_ = 5.26×10^−3^), and *ribosomal protein L10a pseudogene 3, RPL10AP3*, mapped by *rs6990255* (p_FDR_ = 7.54×10^−3^). Another one is consistent with the newest BD GWAS: solute carrier family 25 member 17, *SLC25A17*, mapped by *rs5758064* (p_FDR_= 7.47×10^−3^). The shared genomic loci, candidate independent SNPs, allelic association and novelty for BD were summarized in Supplementary Table S1. The stratified conditional Quantile-Quantile (Q-Q) plots showed SNP enrichment for BDI condition on association with BDII and vice versa (Supplementary Figure S1), suggesting the existence of polygenic overlap.

**Figure 2.**
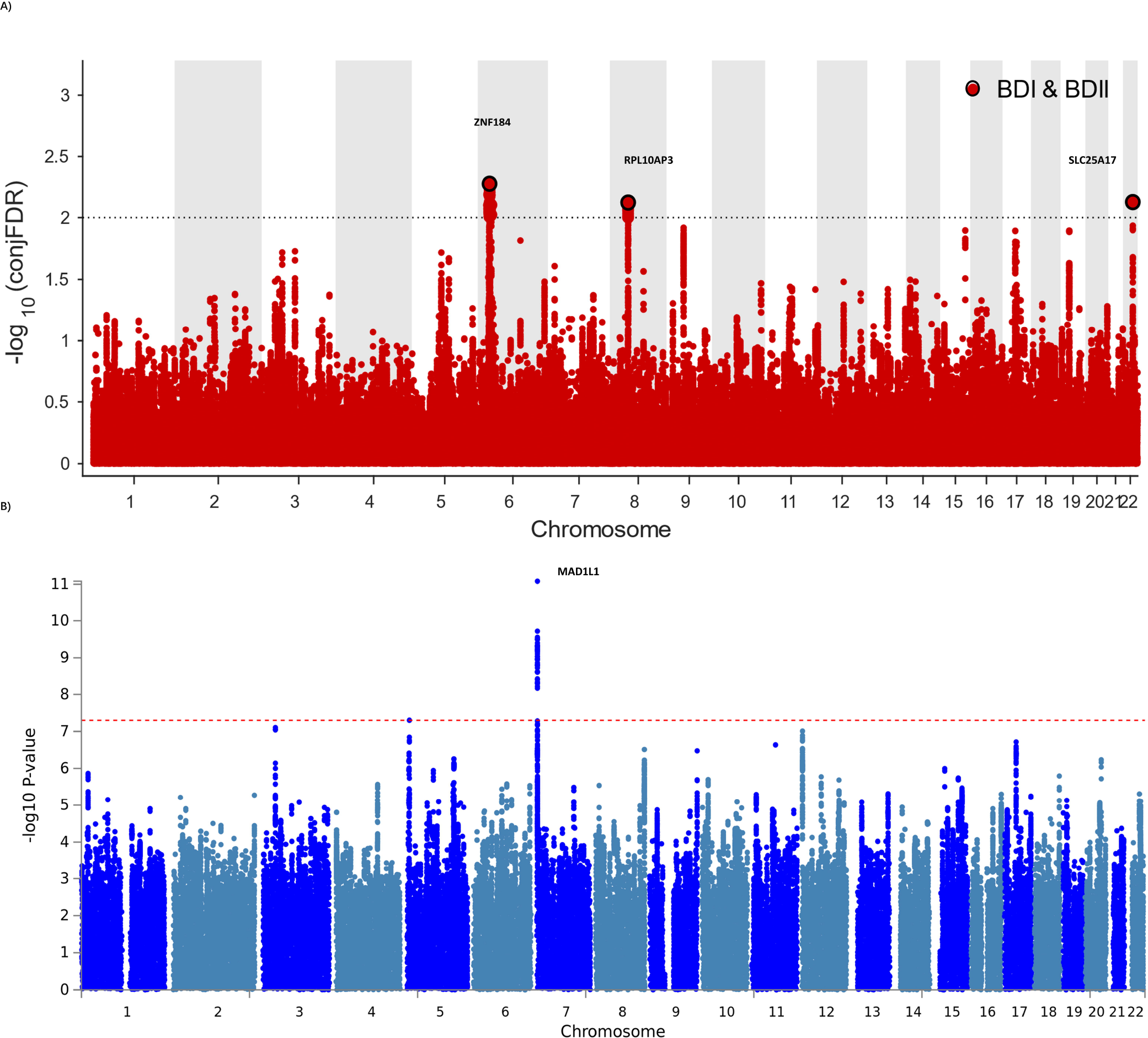
Manhattan plots showing the association statistics for single marker analysis of BD I & II genetic overlap (Figure 2A) and distinctness (Figure 2B). The y-axis shows the GWAS - log10 P-values per SNP across chromosomes 1-22. **A)** represents SNPs with conditional P-value < 1×10^−2^ are shown with large black points. **B)** represents SNPs identified by case-case genome- wide association analysis (CC-GWAS) with p < 5×10^−8^. The figures show the localization of significant loci. Details about the loci are provided in **Table 1** and **Table 2**.

### Genetic distinction of Bipolar type I and II

CC-GWAS analysis was applied to the publicly available summary statistics for BDI and BDII. The only one CC-GWAS BDI versus BDII SNP was *rs12154473*, located on chromosome 7p22.3 (p_OLS_=2.83×10^−8^; p_EXACT_=6.07×10^−5^; Table 2 and Supplementary Table S2 and S3). The Manhattan plot of CC-GWAS results was shown in Figure 2B. The genetic distance between individuals with BDI and BDII is only slightly smaller (0.55) than the case–control distances for BDI (0.62) and BDII (0.62) (Supplementary Figure S2).

In the gene-based analysis, a total of 18,626 genes were tested among them. Nine genes showed distinction between BDI & II after multiple testing (p <0.05/18626=2.68×10^−6^). The top hit was mitotic arrest deficient 1 like 1, *MAD1L1*. Besides, a total of 18,847 or 18,830 genes were tested by FUMA for BDI, BDII, respectively. After Bonferroni correction, 129 genes were significant for BDI, gene *CACNA1C* (p = 2.80×10^−11^), *MAD1L1* (p = 7.56×10^−11^), the same with CC-GWAS, and *TMEM258* (p = 9.48×10^−11^), are the top three genes of BDI. The only significant gene of BDII is *SLIT3* (p = 7.92×10^−9^). Details of genes significantly associated with each phenotype after correction for multiple testing were shown in Supplementary Table S4.

Pathway analyses were next performed for BDI, CC-GWAS and BDII, respectively. 11, 6 and one pathways were significantly enriched by the genes through MAGMA analysis for BDI, CC-GWAS and BDII GWAS summary statistics (P_Bonferroni_ < 0.05). As for BDI, the “neuron part”, “somatodendritic compartment”, “high voltage gated calcium channel activity” and “volted gated calcium channel activity involved in cardiac muscle cell action potential” were gene ontology pathways verified by CC-GWAS and BDI. As for BDII, the only significant pathway was the “Hirsch cellular transformation signature up” (p = 9.84×10^−5^). The mapped pathways and genes were in Supplementary Table S5.

### TWAS analyses in blood and brain regions

SMR was used to prioritize eQTLs playing causal roles in BDI & II via gene expression in blood and brain. 11 in brain (p_SMR_ < 6.61×10^−6^) and 49 in blood (p_SMR_ < 3.19×10^−6^) putative BDI significantly associated genes were identified after multiple testing correction and heterogeneity test. The top loci were NMB and FADS1 for BDI in blood and brain, respectively (Supplementary Figure S3). We did not observe significant results for the smaller sample BDII after multiple testing. The related genomic loci, candidate SNPs and allelic association for BDI were summarized in Supplementary Table S6 and S7.

TWAS was conducted by e-MAGMA and FUSION in 13 brain regions included in GTEx v8. Expression of genes was influenced by trait-associated sites, respectively. E-MAGMA identified 148 loci (p_FDR_<0.05) of BDI. *FADS1* (p_FDR_ = 1.87×10^−7^), *PLEC* (p_FDR_ = 3.10×10^−7^) and *ITIH4* (p_FDR_ = 3.10×10^−7^) were the top ones. These genes encompassed 3 brain regions, including hypothalamus, amygdala, and cerebellum (Supplementary Table S8). Similarly, FUSION indicated 336 genes (p_FDR_<0.05, 9 hits of previous GWAS of BD achieved nominal significance) of BDI (Supplementary Table S9) among all the 13 brain regions.

As for BDII, two genes showed significant association with BDII by e-MAGMA after correction for multiple testing: G protein subunit alpha I 1 on chromosome 7 (*GNAI1*, p_FDR_ = 2.64×10^−7^) and serine protease 16 on chromosome 6 (*PRSS16*, p_FDR_ = 1.40×10^−5^). Both were over-expressed in cerebellum (Supplementary Table S10). 21 genes were significant by FUSION after multiple tests among brain regions without nucleus accumbens (NAc) (Supplementary Table S11). The top hit was *AC005932*.*1* (p_FDR_ = 2.07×10^−4^), a novel nominal significant gene for BDII. The located chromosome regions, correlated brain regions and numbers of association with eQTLs were represented in Figure 3 and Supplementary Table S12.

**Figure 3.**
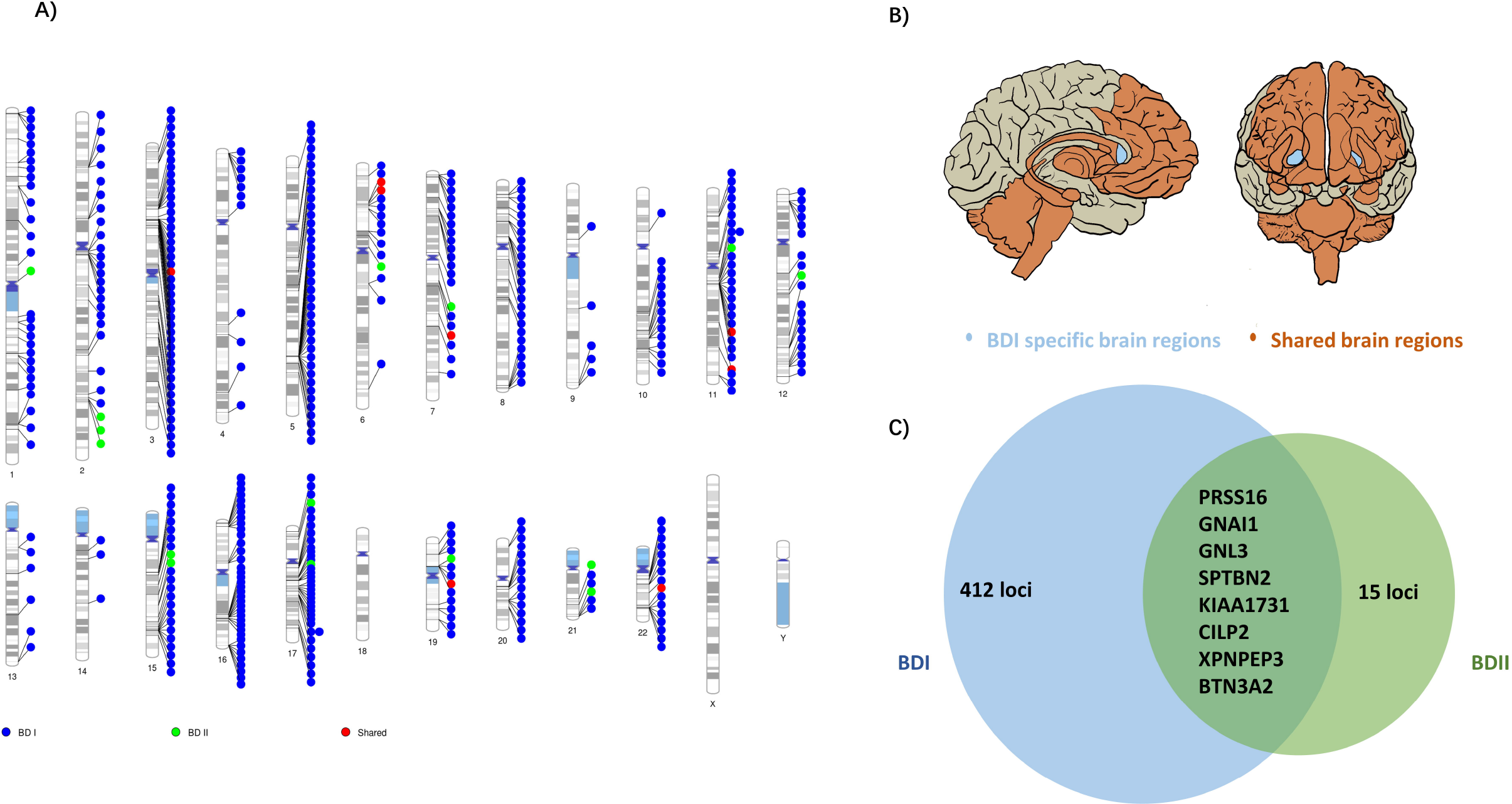
Shared and trait-specific eQTLs of BD I and BD II. **A)** Genomic regions that are specific to BD I (blue points), specific to BD II (green points), and shared (red points). **B)** Associated 13 brain regions with BDI and BDII: cortex, frontal cortex, anterior cingulate cortex, caudate, putamen, hypothalamus, amygdala, hippocampus, substantia nigra, cerebellum, cerebellar hemisphere and spinal cord cervical c-1 (orange area) and BDI specific brain region: nucleus accumbens (light blue area) **C)** Prioritized genes for shared and trait-specific regions. Genes presented reached significance (after FDR correction) in the TWAS test and were identified by two methods (FUSION and e-MAGMA). The numbers represent trait-specific genes identified by at least one method. Details are in Supplementary Table S8-12

### Correlations between BDI & II and response to lithium

Two-sample Mendelian Randomization analysis suggested there might be an effect estimate consistent with increased risk for response to lithium ion (BDI: Weighted median, beta=1.89; standard error (SE), 0.426; p = 9.57×10^−6^), while none for BDII (Inverse weighted regression, beta=3.27; SE, 2.28; P = 0.15). Based on different hypothesis, MR-Egger and MRR-PRESSO displayed opposite results. BDI passed MR-Egger intercept analysis (MR-Egger intercept = 0.41; p = 0.10), while did not pass MR-PRESSO (p_Global_ test < 0.001), indicating existence of horizontal pleiotropy (Supplementary Table S13).

### Genetic overlaps between BDI & II and other traits

Bi-directional MR investigating causal relationships between two BDI & II and traits of interests (SCZ and MDD) was shown in (Supplementary Table S14). GSMR analyses provided evidence that genetically SCZ provided a 0.50-fold and 0.32-fold causality increase in BDI and BDII, respectively. Inversely, MDD increased causality with BDI, BDII by 0.23-fold and 1.11-fold, respectively. In the other direction, BDI provided 0.32-fold causal effect on SCZ (beta=0.32, SE= 0.019, p=3.70×10^−62^), comparing with less effect on MDD (beta=0.043, SE= 0.018, p=1.36×10^−2^). Since the number of SNPs ought to be over 10, p-value threshold was set to be 5×10^−5^ when BDII was computed into clumping as exposure trait. The causal relationship between BDII and SCZ (beta=0.073, SE= 0.008, p= 1.12×10^−21^), were very close to BDII and MDD (beta=0.050, SE= 0.008, p= 7.71××10^−11^). BDI and BDII are presented with causality with each other (Supplementary Table S14)

MiXeR estimated that approximately 7.88 k (SE = 0.26 k) variants influence BDI, which was comparable to the case of SCZ (9.82 k; SE= 0.22 k), lower than that for major depression (21.6 k, SE = 2.64 k) and 19.82 k (SE = 21.12 k) variants influenced BDII. Deficiency of sample size may explain the odd statistics in BDII. MiXeR also revealed a higher polygenicity in BDII and MDD than BDI and SCZ. In BDI and BDII, 7.47 k (SE = 0.29k) and 7.47 k (SE = 1.73k) variants were associated with SCZ, and 5.44 k (SE = 0.59k) and 13.23 k (SE = 5.91k) variants were associated with MDD, respectively. Consistent with LD score regression, MiXeR showed that BDI enjoyed higher genetic correlation with SCZ (rg = 0.70) than MDD (rg = 0.39), and oppositely, BDII possessed higher genetic correlation with MDD (rg = 0.68) than SCZ (rg = 0.61) (Supplementary Figure S4 and Table S15).

## Discussion

The present study is the first comprehensive post-GWAS analysis of BDI & II with latest and largest PGC dataset on bipolar disorder. Different from the original study that aimed to identify novel genes and drug targets using overall BD as primary phenotype^11^, our integrative genomic analyses answered directly to the question remained: what are the shared and distinct genetic components of BD subtypes? Our secondary analyses of the existing dataset focus on the unanswered question in Mullins et al.’s study and generate complementary findings (e.g., novel genes and gene-sets). In this study, we corroborated and expanded evidence from previous clinical and genetic studies that there did exist a partially shared genetic basis between BDI & II and provided further insights into their genetic divergence. When compared to other earlier studies on the same research question (Table S16), our study is innovative from different aspects: 1) larger sample size for both BD I and BD II; 2) more systematic statistical genetics analyses; 3) new biological explanation to the distinction of BD subtypes.

For genetic convergence, three loci were identified shared by BDI & II: *SLC25A17, ZNF184* and *RPL10AP3*. All of these loci were previously reported to be associated with bipolar disorder, depression, ADHD, autism spectrum disorder, or schizophrenia^41-44^, underpinning their contribution to mental disorder risk. *ZNF184* has been reported to be likely associated with subcortical volume^45^, despite the unclear biological function in neurodevelopment. Research for links between mental and physical disorders is also proposed^41, 46^-48

For genetic divergence, notably, *MAD1L1* that was reported to be genome-wide significant in the two previous BD GWAS^49, 50^ including Asian samples, achieved nominal significance distinguishing BDI & II in this study. This gene contributes to cell cycle control through the regulation of mitosis, and has been shown having pleiotropic effect on psychosis and BD before ^15, 51, 52^. Our study suggests that *MAD1L1* may be a genetic marker of BDI, promoting further biological research on it. Moreover, *MRM1, ZNHIT3, DHRS11, GGNBP2* are firstly reported significant in the gene-based test. *SLIT3* was identified to be BDII-specific by gene-based analysis. *SLIT3* has been shown to play a critical role in the formation and maintenance of the nervous system^53^, SCZ^54^, treatment resistant depression^55^ and alcohol dependence^56^, indicating generally shared genetic association among psychiatric disorders.

Enriched gene-sets of BDI were involved in neuronal and postsynaptic compartments, as well as calcium channel activity, triggering presynaptic signaling, which reconfirmed cross-phenotype correlation across BD, SCZ, ASD and cognitive deficiency^16, 57-59^. These pathway processes may indicate BDI primarily represents BD biological features and point deeper research into common biological pathogenesis among mental disorders. As a comparison, the BDII-specific pathway effects generally link with cancer, inflammatory and metabolic diseases^60^, hence suggesting larger cohorts are required to provide mechanic prompt for further research into BDII.

Interestingly, from the integrative Omics analysis, we found *FADS1* as the one of the three top eQTL-associated loci shared by both brain and blood, is presented with opposite directions of effect on gene expression in the two different tissues. This observation also suggests *FADS1* possibly plays a role in the tissue-specific gene regulation of BDI. The possible reason was that *FADS1* is strongly associated with blood cell and lipid and glucose metabolite^61-64^, and thus highly expressed in blood. Brain region-specific eQTL analysis yielded 15 genes specifical for BDII. These eQTLs provide promising candidate genes for subsequent functional experiments, especially *NOS2* (Nitric oxide synthase 2) and *CASP8* (Caspase-8), participate in drug metabolism^65^, despite no correlation to psychosis was yet found. While several of these genes are implicated by genome-wide significant loci, many of them are not the closest gene to the index SNP, highlighting the value of probing underlying molecular mechanisms to prioritize the most likely causal genes in each corresponding locus and moving from genes to functional mechanism.

In addition, the results of this study also suggest that BDI and BDII significantly differ in biosignatures as revealed by gene expression differentiation in functional brain regions and drug response. Gene expression differentiation in NAc might represent an endophenotype of BDI addressing dysfunction of brain circuits. By regulating dopamine release and midbrain dopamine system, NAc contributes to onset of SCZ^66, 67^, especially for delusion and hallucination. It is also a contributor to the pathophysiology of BD, as shown from a postmortem brain analysis^68^. Even though we did not find direct causal relationships between BDI, II and lithium ion, there indeed exists a linkage with lithium response following the guidelines^69^: lithium was first-line to BDI, but not to BDII. Lithium is more effective for patients sharing etiological homogeneity; based on longitudinal stability and familial clustering, lithium response has been suggested to define a distinct genetically based BD nosology^19^. Therefore, biological indicators like treatment response, clinical prognosis and progression of BDI and BDII should be included in genetic analysis, and enable to improve precise clinical decision making. It’s also the RDoC standard that combination of neuroscience research will be helpful for future genetic research, even alter clinical management^70^.

Another interesting finding of this study is the polygenicity and causality of BDI & II with other psychiatric disorders irrespective of genetic correlation^39, 40^. BDI and BDII apparently associated with each other according to the bidirectional causal associations from GSMR and mixed directional overlap from MiXeR in spite that it is confusing to explain whether the causal relationships driven by other covariates^38^. Meanwhile, MiXeR analysis prompted a higher clinical heterogeneity for BDII-MDD pair, when compared with BDI-SCZ pair. Stronger genetic correlation with SCZ inheres in BDI, while a strong association with MDD was seen in BDII. Despite inadequate sample size, consistent results were found from GSMR analysis.

According to our focalization, relationships among these disorders could be that they share continuous genetic structures^71^. BD subtypes may help fill the gap across mental disorders by revealing transdiagnostic biotypes. Insights into such genetic structure may greatly contribute to clinical decision-making in prophylaxis or management of the disorder. This pattern also strongly supports psychiatric disorders could not be regarded as completely independent disease entities. The findings of this study will also trigger larger studies on BDII and other biotypes like psychosis bipolar disorder and cyclothymia, because current BD GWAS mainly reflected the genetic characteristics of BDI, the majority of overall BD cases. Although a large sample size of GWAS is always important in nowadays genomic studies, the statistical power will be greatly reduced when there is nonnegligible clinical heterogeneity caused by the classification system within disease phenotype^72^. Therefore, as RDoC emphasized, large-scale transdiagnostic investigations are urgently needed to untangle whether impairment or symptoms can be regarded subtype-specific, and so do multi-omics analysis^73^.

One of the potential limitations of our study is that the population imbalance of BDII GWAS summary statistics may be susceptible to bias. We were restricted to find more loci in each level of our study compared with its relatively higher heritability to MDD. Further collection and analysis should be emphasized on BDII. Another limitation is that we failed to achieve individual genotypes, leading to the incompleteness of some important analyses, such as polygenic risk score (PRS) calculation, and so on. Finally, our study only obtained GWAS summary statistics of BDI and BDII, lacking data from other BD subtypes.

In summary, genetic evidence deepens our understanding of the biological etiology of BD and prioritizes a set of candidate genes distinguishing BDI & II for functional follow-up experiments and indicates a spectrum connecting psychiatric disorders, which enable better ways to optimize nosology and precise treatments in psychiatry.

## Data Availability

All data produced in the present study are available upon reasonable request to the authors. All data produced in the present work are contained in the manuscript. All data produced are available online at the Psychiatric Genomics Consortium Bipolar Disorder Working Group (PGC3 BD) and National Human Genome Research Institute-European Bioinformatics Institute (NHGRI-EBI) GWAS Catalog20 (https://www.ebi.ac.uk/) and PGC website.

https://www.med.unc.edu/pgc/

https://www.ebi.ac.uk/

## Acknowledgments

This study was financially supported by the National Nature Science Foundation of China (Grant NO.81771446 to QW, NO.82171499 to QW and 82071524 to ML), Science and Technology Project of Sichuan Province (2021YJ0238 to ML). HG is supported by Mentored Scientist Grant in Henry Ford Hospital.

## Table Legend

**Table 1. Conjunction FDR; pleiotropic loci in BD type I (BD I) and BD type II (BD II) (BDI&BDII) at conjFDR < 0.01.** Independent complex or single gene loci (r^2^ < 0.2) with SNP(s) with a conjunctional FDR (conjFDR) < 0.05 in (BDI) and (BDII). All SNPs with a conjFDR value,0.05 (bidirectional association) and association with BDI & BDII are listed and sorted in each LD block. We defined the most significant SNP in each LD block based on the minimum conjFDR. Chromosome (CHR), minor allele (A1) and major allele (A2), Z-scores for each pleiotropic locus are provided. All data were first corrected for genomic inflation. Locus name is based on exonic lead SNPs. Remaining locus name is based on the nearest gene and do not refer to any inferred biological function. Details are in Supplementary Table S1.

* Same locus identified in previous BD genome-wide association studies

**Table 2. Summary of CC-GWAS results for BDI&BDII.** For the CC-GWAS-specific locus, the lead CC-GWAS SNP and its chromosome, physical position, the locus name, the respective case– control effect sizes and P values and the CC-GWAS OLS and Exact case–case effect size, standard error(se) and P values. Details are in Supplementary Table S3.

