## Supplementary Figures for "Patterns of convergence and divergence between bipolar disorder type I and type II: evidence from integrative genomic analyses"

**Figure S1**

**Figure S2**

**Figure S3**

**Figure S4**

**Figure S5**

**Figure S6**

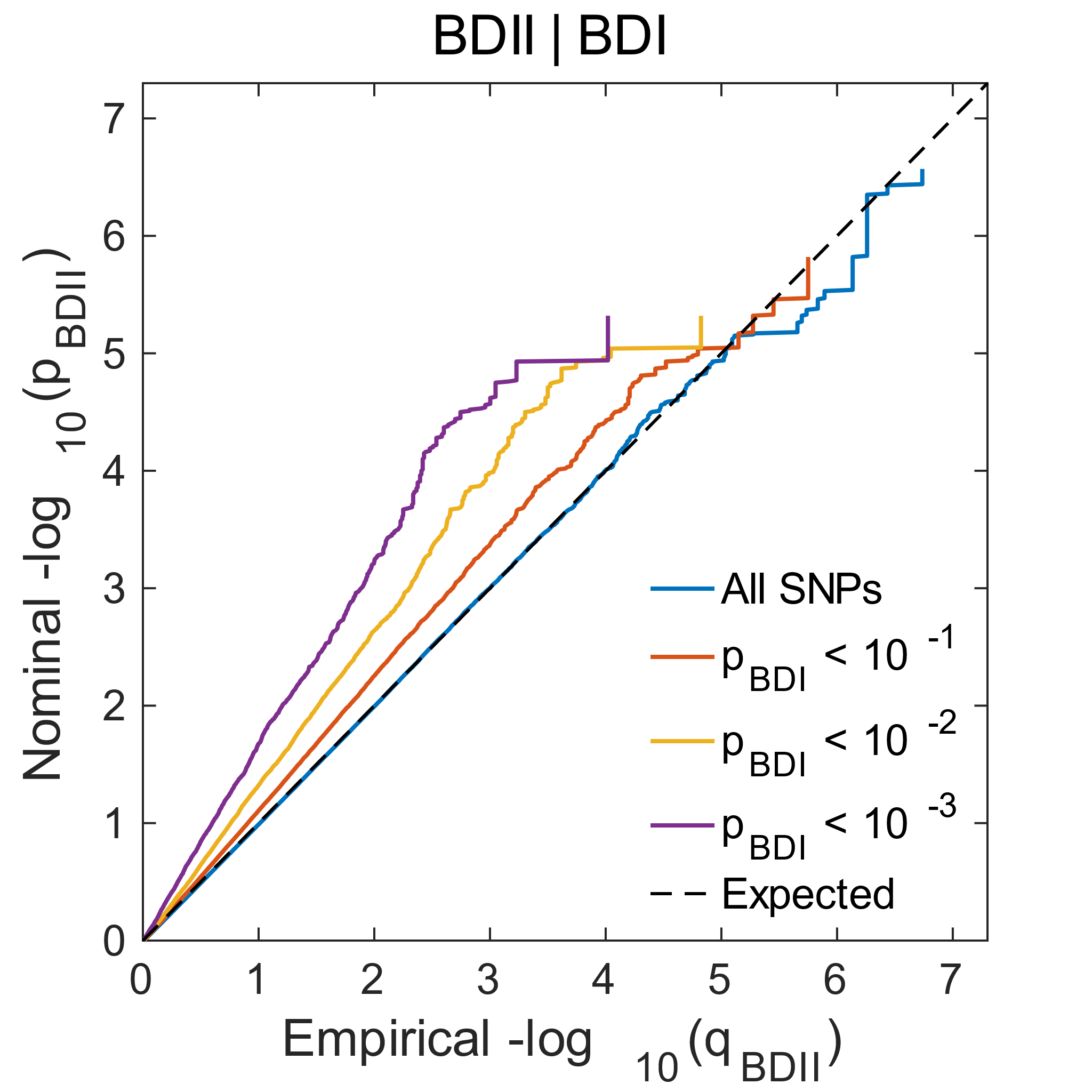

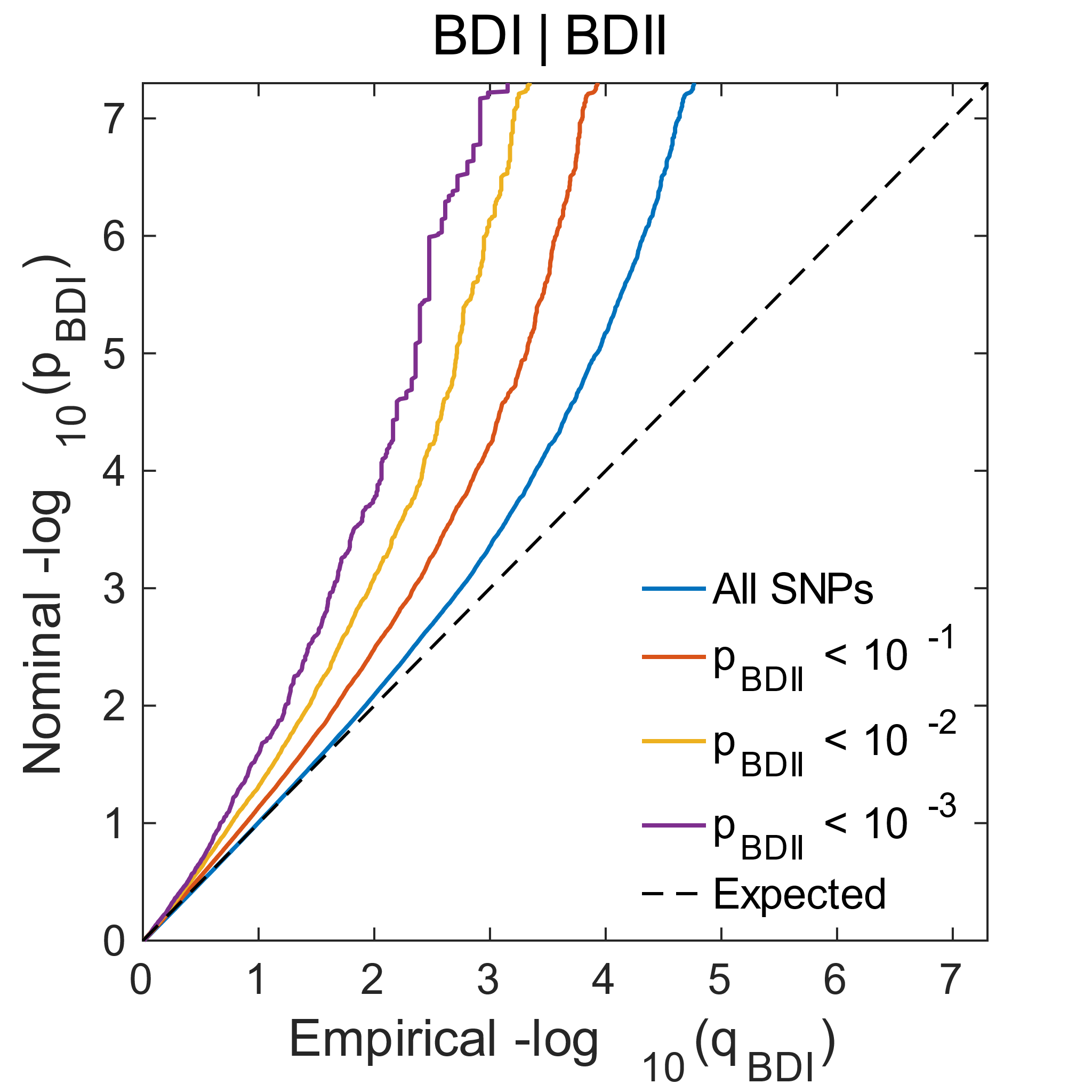

**A)**

**B)**

**Supplementary Figure S1. Polygenic overlap between BD I and BD II. Conditional Q-Q plots of nominal versus empirical −log_10_ p-values (corrected for inflation) in BD I below the standard GWAS threshold of p<5×10^−8^ as a function of significance of association with BD II, at the level of p ⩽ 0.1, p ⩽ 0.01, p ⩽ 0.001, respectively, and vice versa. The blue lines indicate all SNPs. The dashed lines indicate the null hypothesis.**

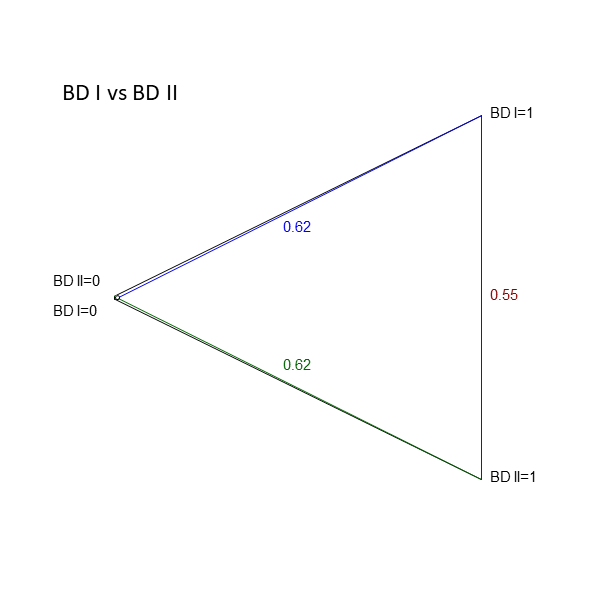

**Supplementary Figure S2. Genetic distance between cases and/or controls of BD I & II. Genetic distances are displayed as** $\sqrt{\boldsymbol{m*F(ST,causal)}}$**, derived based on the respective population prevalences, SNP-based heritabilities and genetic correlations. The genetic distance between individuals with BD I and II (BD I cases and BD II cases) is only slightly smaller (**$\sqrt{\boldsymbol{m*F(ST,causal, A}\boldsymbol{1}\boldsymbol{B}\boldsymbol{1)}}$**=0.55) than the case–control distances for BD I (**$\sqrt{\boldsymbol{m*F(ST,causal, A}\boldsymbol{1}\boldsymbol{A}\boldsymbol{0)}}$**=0.62) and BD II (**$\sqrt{\boldsymbol{m*F(ST,causal,B}\boldsymbol{1}\boldsymbol{B}\boldsymbol{0)}}$**=0.62), because of little imbalance between BDI and BDII prevalence. Also, BD I & II are both subtypes of BD, sharing similar genetic structures. Also, BD I & II are both subtypes of BD, sharing similar genetic structures.**

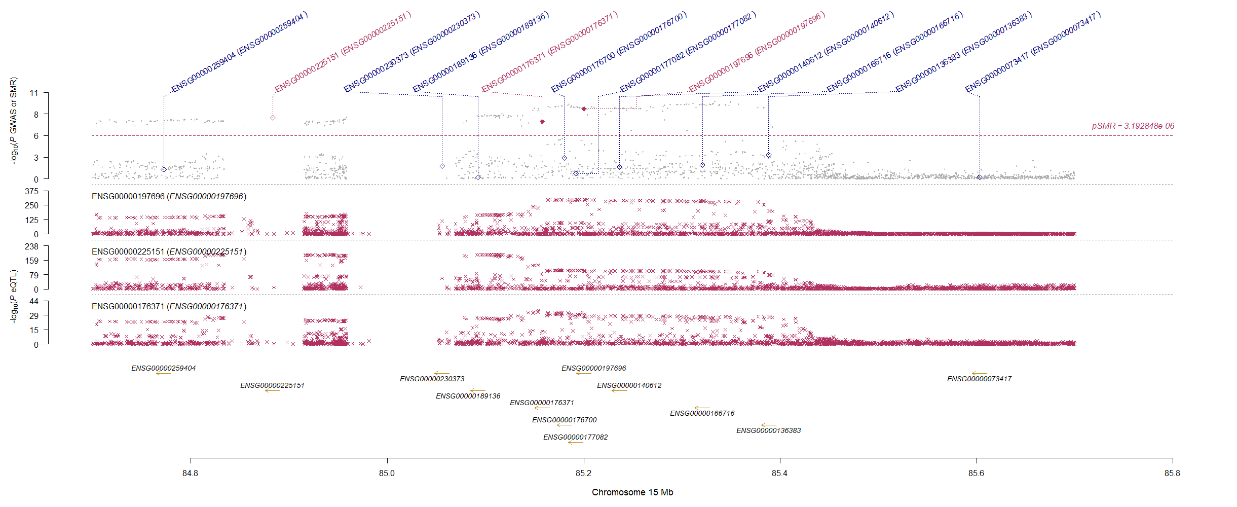

**Supplementary Figure S3. Prioritizing genes at the most significant loci for BDI in blood (A) and brain (B). Shown are results detected by Summary-data-based Mendelian Randomization (SMR) analysis using summary data from PGC3 BDI & II and eQTLGen Consortium blood data (A) and GTEx brain data (B). Top plot, grey dots represent the P values for SNPs from GWAS, diamonds represent the P values for probes from the SMR(A: p= 0.05/15659=3.19 × 10^−6^, B: p= 0.05/7443=6.72 × 10^−6^). Bottom plot, P values from eQTL analysis for NMB (the top probe in blood)/GOLGA2P7/ZSCAN2 and FADS1(the top probe in brain). Probes that passed the HEIDI test (PHEIDI ≥ 0.05) are highlighted in red.**

**A)**

**B)**

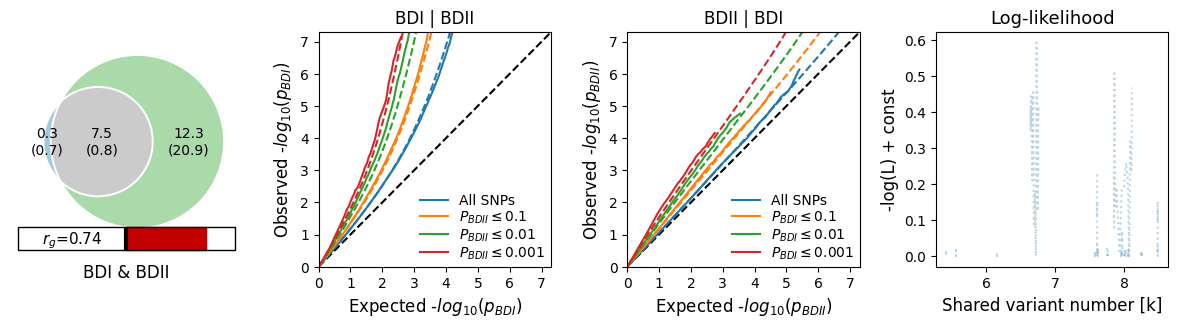

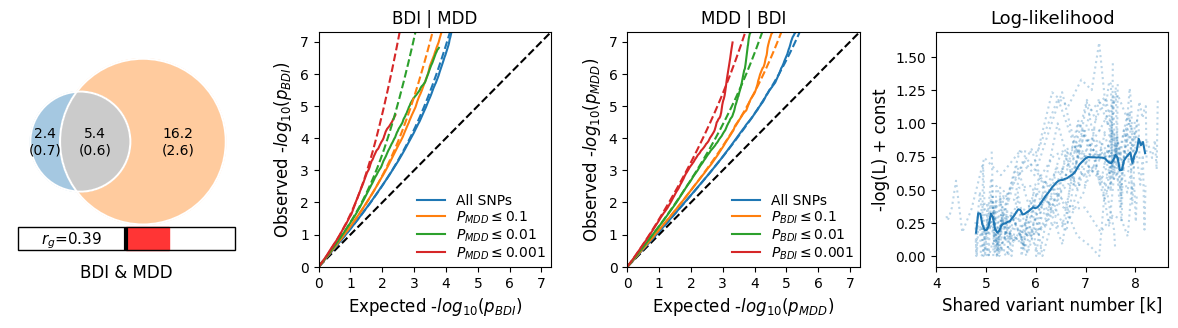

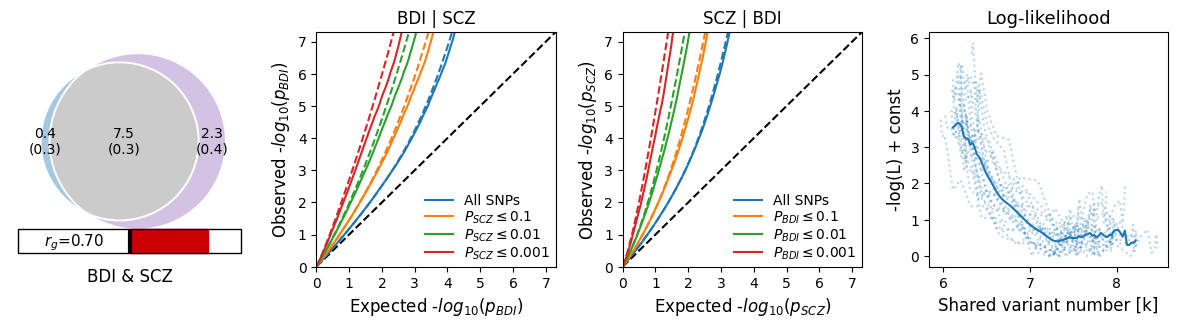

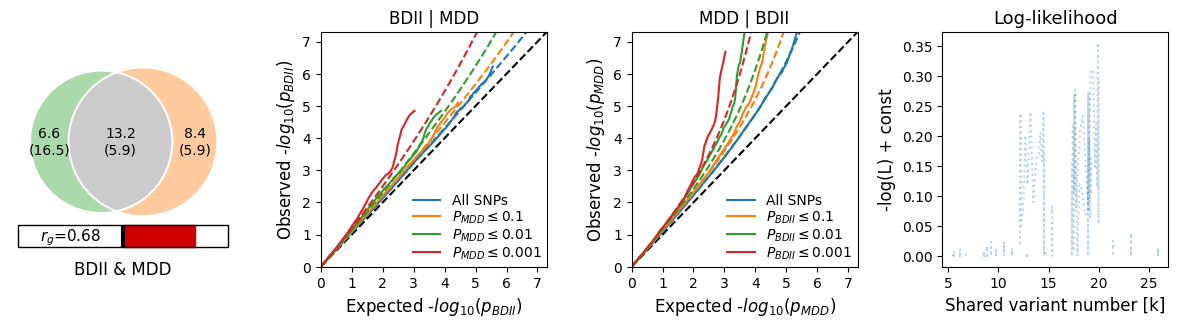

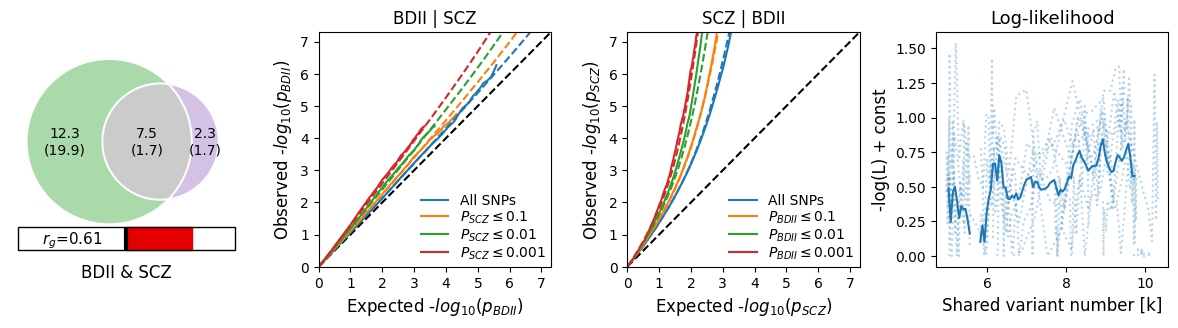

**A)**

**B)**

**C)**

**D)**

**E)**

**Supplementary Figure S4. Venn diagrams of unique and shared polygenic overlap at the causal level (A-E). (gray) between BDI (blue), BDII (green), SCZ (purple), and MDD (orange). The numbers indicate the estimated quantity of causal variants (in thousands) per component, explaining 90% of SNP heritability in each phenotype, followed by the standard error. The size of the circles reflects the degree of polygenicity. Conditional Q–Q plots of observed versus expected −log_10_ *p*-values in the primary trait as a function of significance of association with a secondary trait at the level of *p* ≤ 0.1 (orange lines), *p* ≤ 0.01 (green lines), *p* ≤ 0.001 (red lines). Blue line indicates all SNPs. Dotted lines in blue, orange, green, and red indicate model predictions for each stratum. Black dotted line is the expected Q–Q plot under null (no SNPs associated with the phenotype).**

**Supplementary Figure S5. Venn diagrams of shared and unique eQTL numbers among BDI, BDII, SCZ and MDD by e-MAGMA (A), FUSION (B) and both (C). Trait-specific eQTLs are highlighted in blue, orange, purple and green. Trait-shared eQTLs are highlighted in dark.**

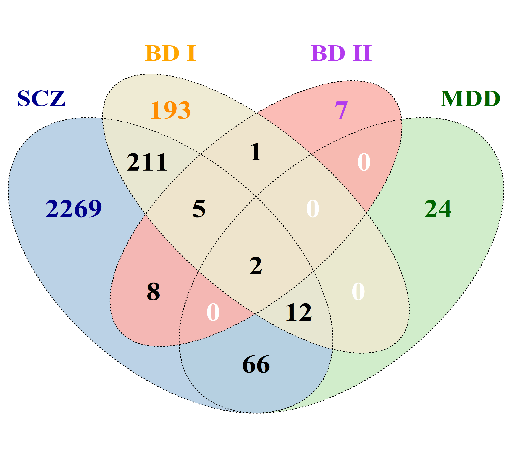

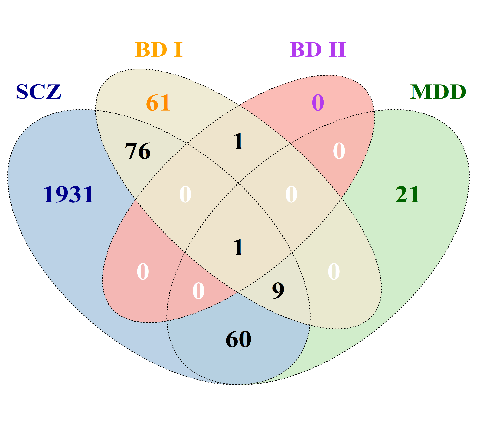

**A)**

**B)**

**C)**

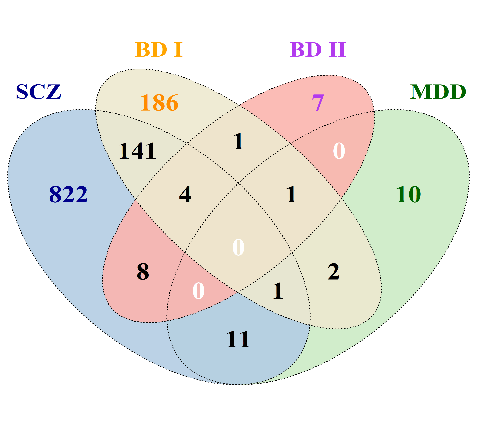

**E-MAGMA**

**FUSION**

**E-MAGMA & FUSION**

**A)**

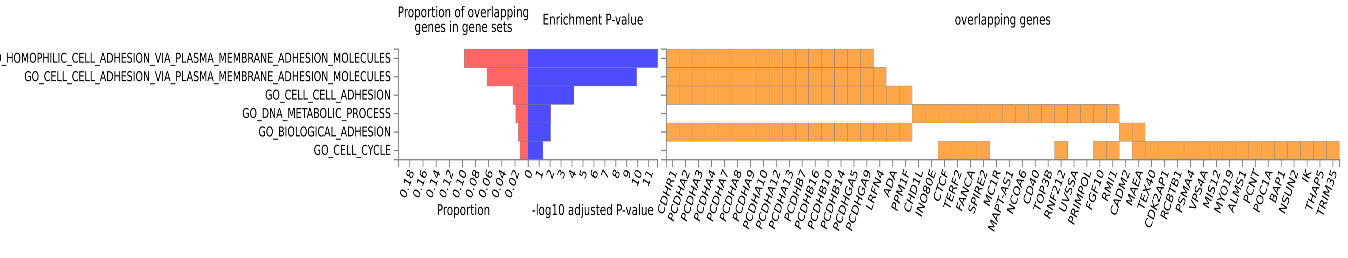

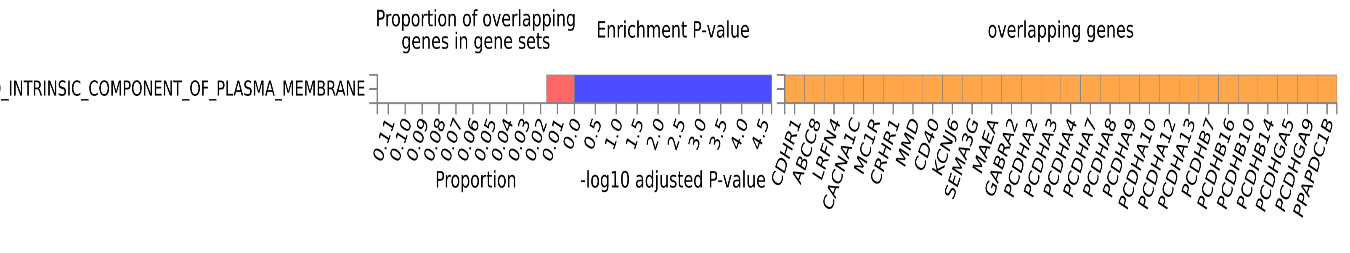

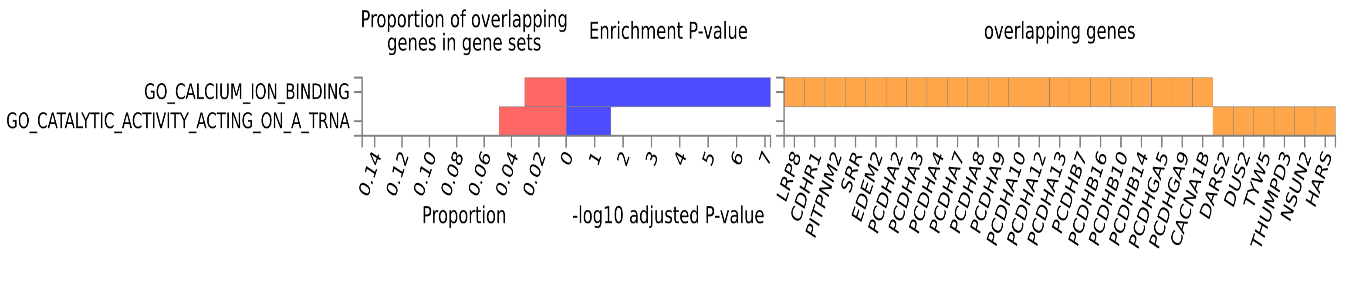

**Supplementary Figure S6. BDI and SCZ shared genes enriched in Gene ontology (GO) pathways. Shown are gene sets with adjusted P-value < 0.05 in GO biological processes(A), cellular components(B) and molecular functions(C).**

**C)**

**B)**
